# An Open-Label, Interventional, Prospective, Real-World Evidence Study to Evaluate a Multimodal Wound Matrix in Patients with Refractory Wounds

**DOI:** 10.1101/2024.10.01.24314439

**Authors:** Yadwinder Dhillon, Gerit Mulder, Keyur D. Patel, Luis Moya, Gerard Boghossian, David Swain, Robert B. McLafferty, Kelly Perez, Jessica Nguyen, Natalie Wilkinson, Jessica Arragon, Lilia Contreras, Donna Geiger, Ryan Cummings, Brenda LaVigne, Desmond P. Bell, Suzanne J. Bakewell

## Abstract

**Objective:** The objective of this open-label, interventional, prospective clinical study was to evaluate the effectiveness of a multimodal wound matrix (MWM), formulated for the treatment of chronic, non-healing wounds that had failed prior therapies. The overall response rate was the proportion of patients that had greater than 40% reduction in size after 4 weeks of treatment. Secondary objectives included the percentage area reduction (PAR) after 4 and 12 weeks, incidence of ulcers closing, and changes in quality of life.

**Approach:** An open-label, interventional, prospective cohort, real-world evidence study was conducted at 8 US wound care sites. Criteria included chronic non-healing wounds of multiple etiologies in patients with extensive comorbidities. Results were compared to data from the US Wound Registry.

**Results:** 111 patients entered the screening phase and 63 were treated. 54 wounds were eligible for the dataset that included 18 DFUs, 19 VLUs, 2 pressure injuries, 1 surgical, 1 lower extremity wound, and 12 unclassified etiologies. The objective response rate was 42%. The 4-week PAR was 34%. The 12-week PAR was 66%. 18 wounds closed by week 12.

**Innovation:** MWM is a formulation technology developed to address the major obstacles that prevent healing. Results were evaluated in a patient population with extensive comorbidities who had failed prior treatments and would be generally excluded from controlled trials.

**Conclusion:** The results from this study support the contention that the multimodal wound matrix achieves substantial clinical improvement in a complex patient population not enrolled in clinical trials and demonstrates an advancement in wound management.

## INTRODUCTION

Chronic, non-healing wounds represent significant morbidity and mortality, especially to the elderly and those with coexisting conditions or compromised immune systems.^1-4^ Refractory wounds are chronic wounds that do not respond to treatment, including evidence-based modalities. A wound is considered chronic when it doesn’t progress through an orderly and timely healing process. The healthcare cost of these chronic wounds can add up to 2-3% of the healthcare budgets in developed countries.^5^

We conducted a real-world study of refractory wounds treated with a multimodal wound matrix (MWM) to more accurately reflect the needs of high-risk patients with multiple comorbidities who are typically managed in wound centers and other clinical settings. The aim was to evaluate clinical improvement in previously unresponsive wounds among a high-risk patient population that would be excluded from controlled trials, given that the clinical outcomes may more accurately translate into a greater potential impact on this population’s long-term outcomes. Our main objectives were (i) to evaluate the potential effectiveness of MWM in the treatment of chronic nonhealing wounds or ulcers across a range of etiologies in wound clinics, (ii) to evaluate the impact on the healing trajectory, specifically moving stalled wounds towards healing, and (iii) to evaluate the percent area reduction (PAR) of wounds after 4 weeks of MWM treatment. Secondary objectives were (i) change in quality of life (QoL), (ii) enhanced activities in daily living, (iii) time to maximum closure or complete closure, and (iv) incidence of healing at week 12 as end of study (EOS). Our Objective Response Rate (ORR) was defined as the proportion of subjects that had at least a 40% reduction after 4 weeks of treatment.

MWM’s formulation was designed specifically to assist the body with the healing cascade and to address the characteristics common to all chronic wounds, regardless of etiology. These characteristics include edema, biofilm colonization, and decreased localized perfusion. MWM is comprised of components derived from marine and plant sources, including omega fatty acids, medium and long chain fatty acids, and cold-water fish peptides, whose benefits are relevant to wound healing.^6^ Specifically designed both chemically and physically to target the major hurdles obstructing the healing process, MWM treatment aims to convert these wounds into a healing trajectory.^6^ This study was conducted to evaluate the effectiveness and efficacy of MWM’s formulation on the wounds it was designed to treat.

## MATERIALS & METHODS

### Patient Population

This study (NCT05921292) was an open-label, interventional prospective real-world evidence study that was conducted from April 2022 through April 2024. It was designed to evaluate the effect of a multimodal wound matrix (MWM) in a cohort of patients with refractory wounds. A refractory wound was defined as a wound that had not responded to treatment for more than 8 weeks, and after a minimum of 2 weeks of standard care in a specialty wound clinic. The intent of the study design was to include patients that are commonly treated daily in wound care clinics. The protocol included a broad inclusion criteria and narrow exclusion criteria in an attempt to eliminate the bias that can experienced in controlled trials. The aim was to enroll approximately 111 subjects with an expectation of 100 completers.

Ethical approval for the study was given by Salus Institutional Review Board (OM-CTP-002), Austin, TX, USA, and by Oregon Health and Science University (OHSU) Institutional Review Board ((STUDY00024987), Portland, OR, USA. The study was conducted in accordance with the principles consistent with the Declaration of Helsinki, Good Clinical Practice, applicable regulatory requirements, and the Belmont Principles of respect for persons, beneficence, and justice. All patients provided written informed consent to participate in the study.

The clinical sites in this study were located in the US and included hospital outpatient wound care centers, wound and research offices, and physician offices. Subjects were selected by the Principal Investigators (PIs) from their respective internal patient populations within their clinical sites of service. Patients treated for at least 2 months with no significant response were identified as potential subjects for the study. Eligibility criteria included active smokers, BMI<65, and opioid users. Patients on dialysis or with wounds less than 2cm^2^ in size were excluded. Following the inclusion/exclusion criteria, subjects underwent the screening visit as part of a 2-week run-in period (Figure 1).

**Figure 1.**
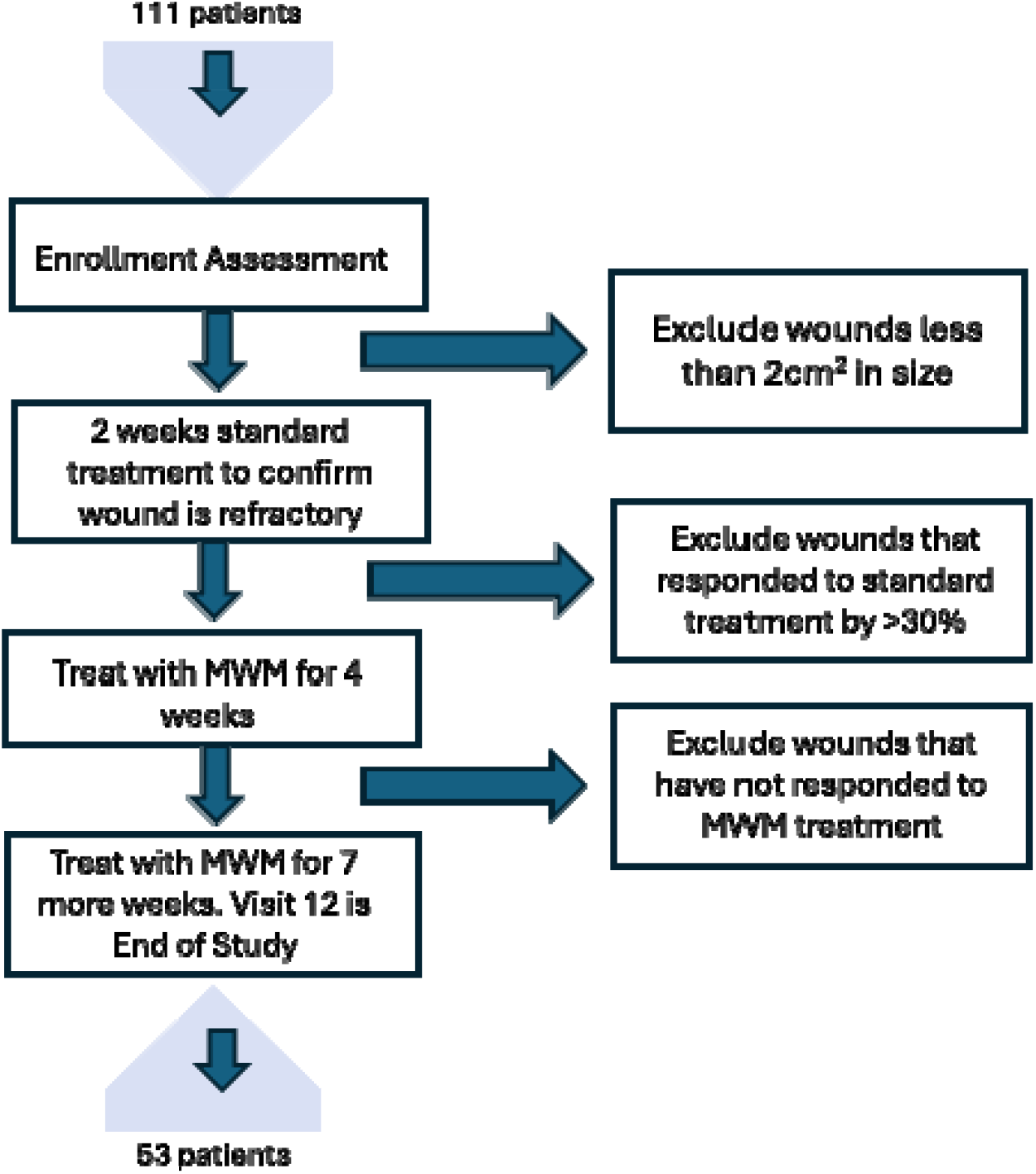
Schematic of the trial design depicting the enrollment assessment, the 2-week screening period, the treatment periods after the first 4 weeks and at the end of study visit. Reasons for wound exclusions are shown at each stage (MWM: Multimodal Wound Matrix)

Assessments were taken at the initial screening visit (SV1) and included a medical history, physical examination, vital signs, treatment history of the chronic wound/ulcer, and target wound determination and assessment for extrudate signs and symptoms and infection. Quality of life (QoL) and activities in daily living (ADL), pain assessment on a visual analog scale (VAS), and percentage body surface area (BSA) involvement were also collected. Additional inclusion criteria for venous leg ulcers required vascular assessment to confirm adequate perfusion, confirmation of venous disease, biopsy performed on ulcers older than 6 months, and compression bandaging. Additional inclusion criteria for diabetic foot ulcers included confirmation of Type I or Type II diabetes, arterial testing, known HbA1c of ≤12% within 6 months of the study, and off-loading.

The screening visit included digital imaging and measuring to confirm that wounds were refractory and non-responsive after the 2 weeks of standard treatment. Wounds were cleaned, debrided as necessary, and dressed. Diabetic foot ulcers (DFU) received off-loading and venous leg ulcers (VLU) received compression wraps during the screening period. Ulcers that responded to this treatment and reduced in size by more than 30% over the 2-week screening period were recorded as non-refractory and did not continue in the study. Eligible patients were treated once weekly with MWM. Patients that missed more than one treatment visit were determine non-compliant and excluded from the data set. Wounds were evaluated and measured after 4 weeks of treatment at treatment visit 5 (TV5). At that time, if there was a positive response to treatment with MWM, the PI could choose to continue for 7 more weeks of treatment with MWM. This decision did not require a reduction in wound size but could be determined by the PIs judgement on the wound’s response to treatment based on for example, status of granulation tissue, increased perfusion, tissue color, or other signs of a healing response to the treatment. Visit 12 was the end of study (EOS) after 11 treatments and final measurements were compared to baseline to determine the percent area reduction (PAR). 100% PAR and closure of the wounds was determined as 100% re-epithelialization with no drainage or exudate present. There was no follow-up with the patient after closure.

### Treatment Protocol

The subjects were treated weekly with MWM (OCM™, Omeza® LLC, Sarasota, FL) following the manufacturer’s Instructions for Use (IFU). The wound was cleaned and debrided per Investigator’s discretion before MWM was applied in strips to the wound bed. Wound images were captured, traced and reviewed at each visit. Approximately 3-5 minutes after the application, the MWM was spread evenly throughout the wound. A non-adherent primary dressing was applied to the wound following the application of MWM.

### Safety

Assessment of adverse events (AEs) and serious adverse events (SAEs), treatment-related and non-treatment-related were monitored until resolved. Concomitant medication changes were also reviewed at each visit. Twenty-four patients experienced a total of 43 adverse events (AEs) and 7 serious adverse events (SAEs). The SAEs were unrelated to the study drug. There were 8 AEs considered probable, 3 possible, and 1 definitively linked to the investigational product. Thirty-one AEs were not linked to the product.

### Data Imaging

Swift® mobile application was used diagnostically to capture wound images, wound measurements and visit date at every visit. Swift® follows industry best practices and complies with all applicable industry standards such as HIPAA and FDA standards. Data and images were stored securely in AWS cloud with daily RDS backups. Photographs were examined to ensure elimination of patient identification. Swift measurements of the wounds were compared at treatment visit 1, treatment visit 5 after 4 weeks of treatment, and at week 12 which was end of study (EOS) visit after 11 weeks of treatment. If wound closed prior to week 12, that visit became end of study.

### Statistics

All analyses of data from this study were descriptive (without p value generation) as the study was not powered for inferential analyses and no formal hypothesis testing was performed. As a single cohort study, variables of interest were compared by wound type. Average, median and the range of wound sizes captured by Swift imaging were evaluated by each wound type.

## RESULTS

One hundred eleven patients signed informed consent forms (ICFs) from eight wound care sites across the United States over a 20-month period from April 2022 through January 2024. Following the 2-week screening period, 12% of patients were discontinued in the trial since their wounds decreased in size greater than 30%, and 23% screen failed for wounds being too small. 64 wounds were treated with MWM. Eleven treated wounds were ineligible for data analysis as 7 wounds were under 2cm^2^ at study start, 2 wounds had increased greater than 30% after the screening phase, 1 patient had 2 wounds with measurements recorded inconsistently, and 1 patient did not have an initial measurement recorded.

The study population (n=53) included 34 males and 19 females (Figure 2a, 2b). Patient ages ranged from 35-100 years (Table 1) with an average of 64.3 years (Median: 63 years). All patients over 65 presented with 2 or more comorbidities (range: 2-26 per patient). 17/25 patients presented with 5 or more comorbidities. In patients under 65, the comorbidity count ranged from 2 to 23 with 12/28 having 5 or more comorbidities. Body Mass Index (BMI) ranged from 15.94 – 52.94 with an average of 33.5kg/m^2^ (Median: 31.8 kg/m^2^).

**Figure 2.**
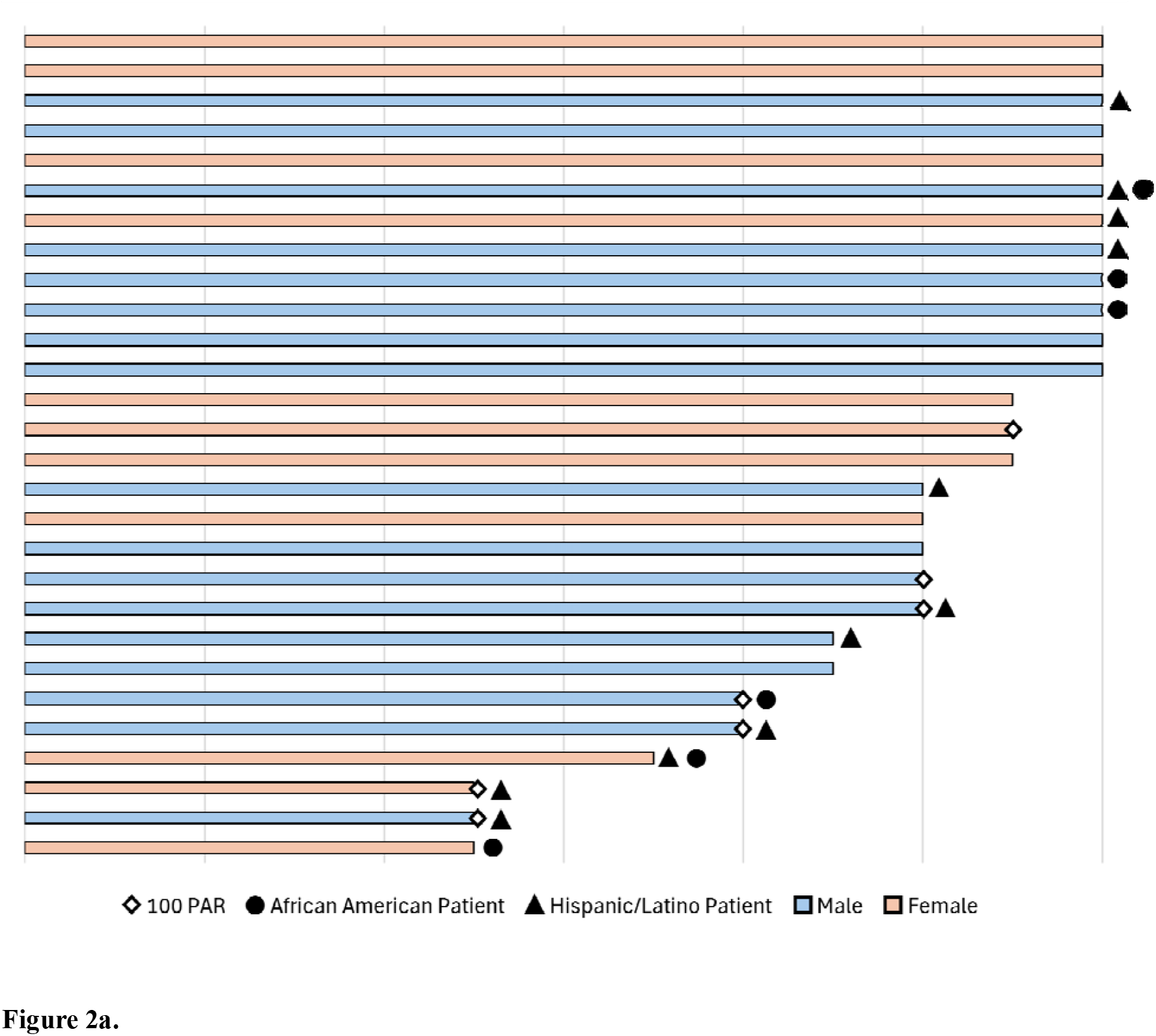
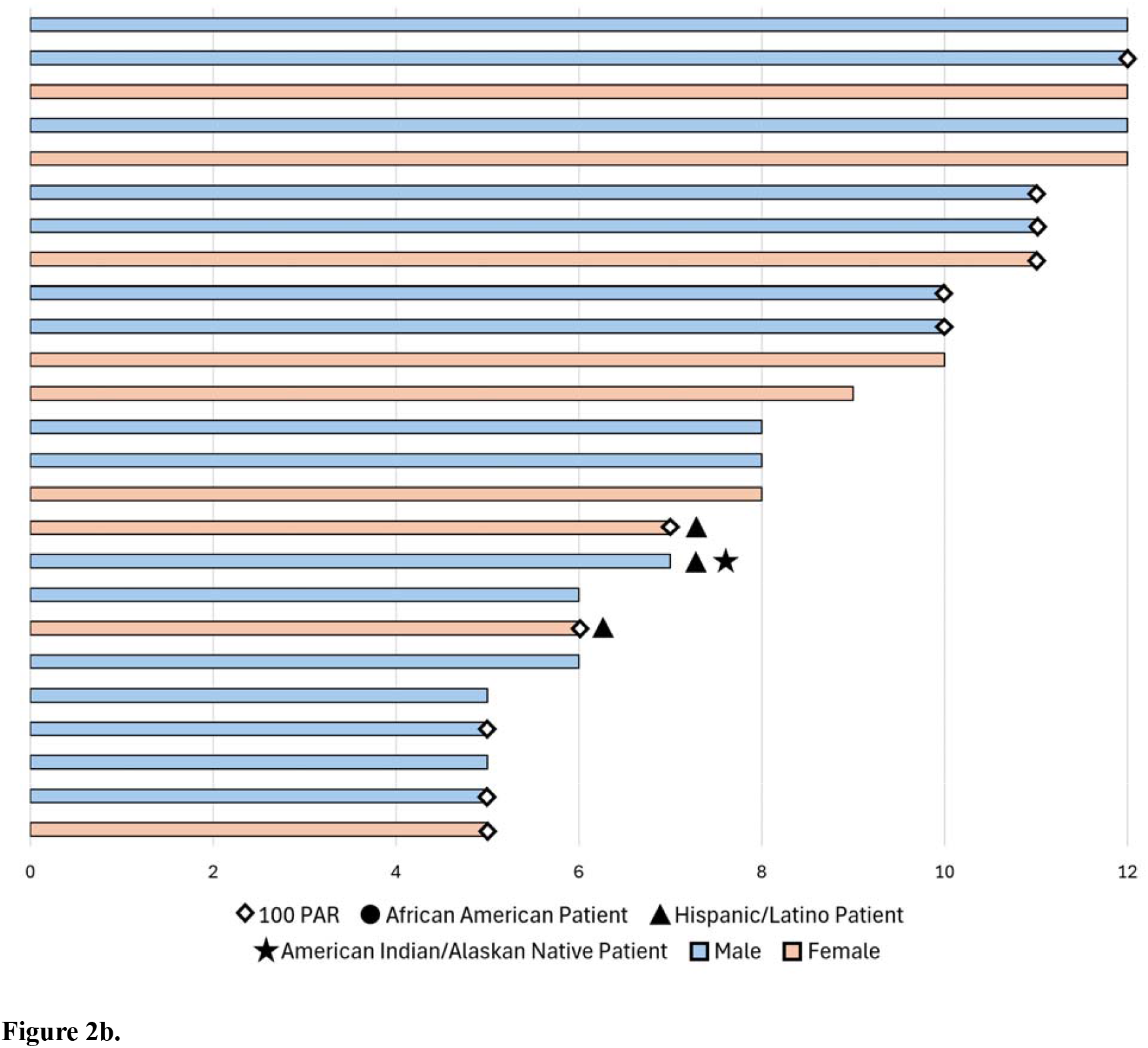
Swimmer plots showing patients demographics, gender, length of treatment time and patients with 100% area reduction, grouped by age range. (a) Patients under 65 years of age (b) Patients over 65 years of age

**TABLE 1.**
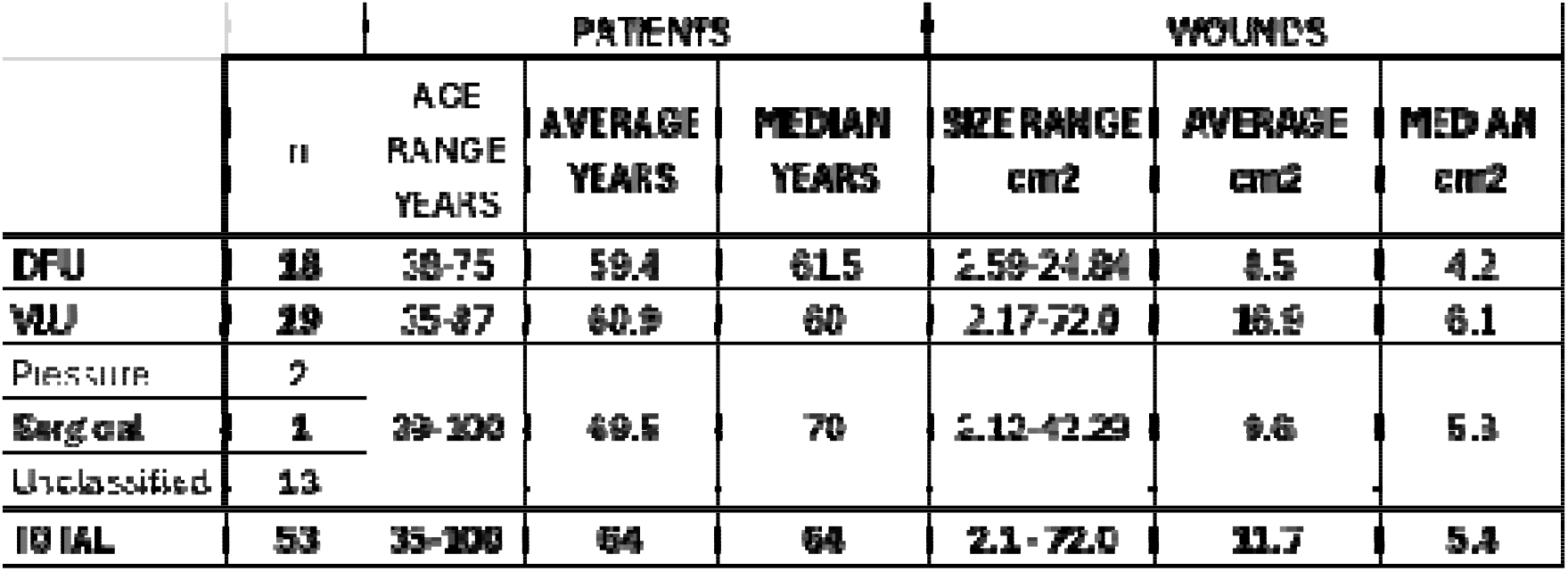
Patient ages and wound sizes.

The wounds treated included 18 DFUs, 19 VLUs, 2 pressure injuries, 1 surgical wound, 1 lower extremity ulcer, and 12 unclassified ulcers. Wound sizes ranged from 2.1cm^2^ to 72.0cm^2^ with an average of 11.8cm^2^ (median: 5.9cm^2^) (Table 1). The age of the wounds ranged from 10 to 304 weeks with an average of 38 weeks (median:22 weeks). Six wounds were older than a year with 3 wounds older than 3 years.

The overall 4-week PAR was 34% and the overall 12-week PAR was 66% (Table 2). The objective response rate was 42% with 22 of the 53 wounds decreasing in wound size by more than 40% after 4 weeks of treatment. Four wounds closed by 100% after 4 weeks of MWM treatment. 34% (18/53) of the wounds saw 100% re-epithelialization by week 12. 32% (16/53) of wounds did not respond at all to treatment, 8 of which were labeled ‘unclassified’. 55% (29/53) of the wounds decreased by more than 40% by week 12.

**TABLE 2.**
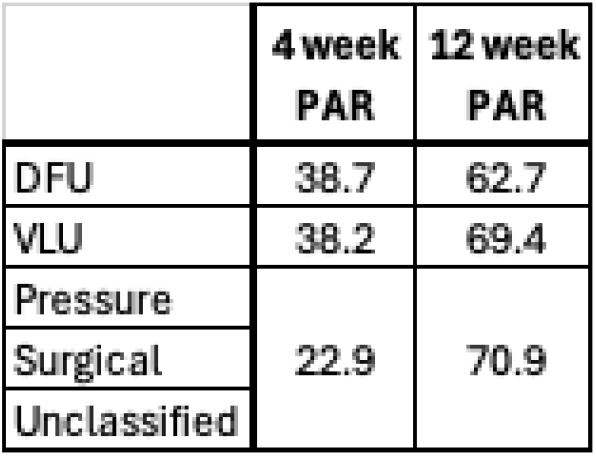
The percentage area reduction (PAR) after 4 weeks of treatment and at 12 weeks presented by wound type.

### Subpopulations

Due to the complexity of the comorbidities of the patient population, the wounds were grouped by the following wound types.

### Diabetic Foot Ulcers

18 patients (17 male, 1 female) with the average age of 59.4 (Median: 61.5) and ranging from 38-75 years were treated with MWM. The ulcer sizes ranged from 2.59-24.84cm^2^ with an average size of 8.47cm^2^ (median: 4.21cm^2^). The age of the ulcers ranged from 10 to 34 weeks, with the average age at 20 weeks (median: 18). The 4-week average PAR was 39% (Figure 3a). 13 of the ulcers were treated up to 12 weeks and had an average PAR of 63% (Figure 3a) with 6 wounds closing completely.

**Figure 3.**
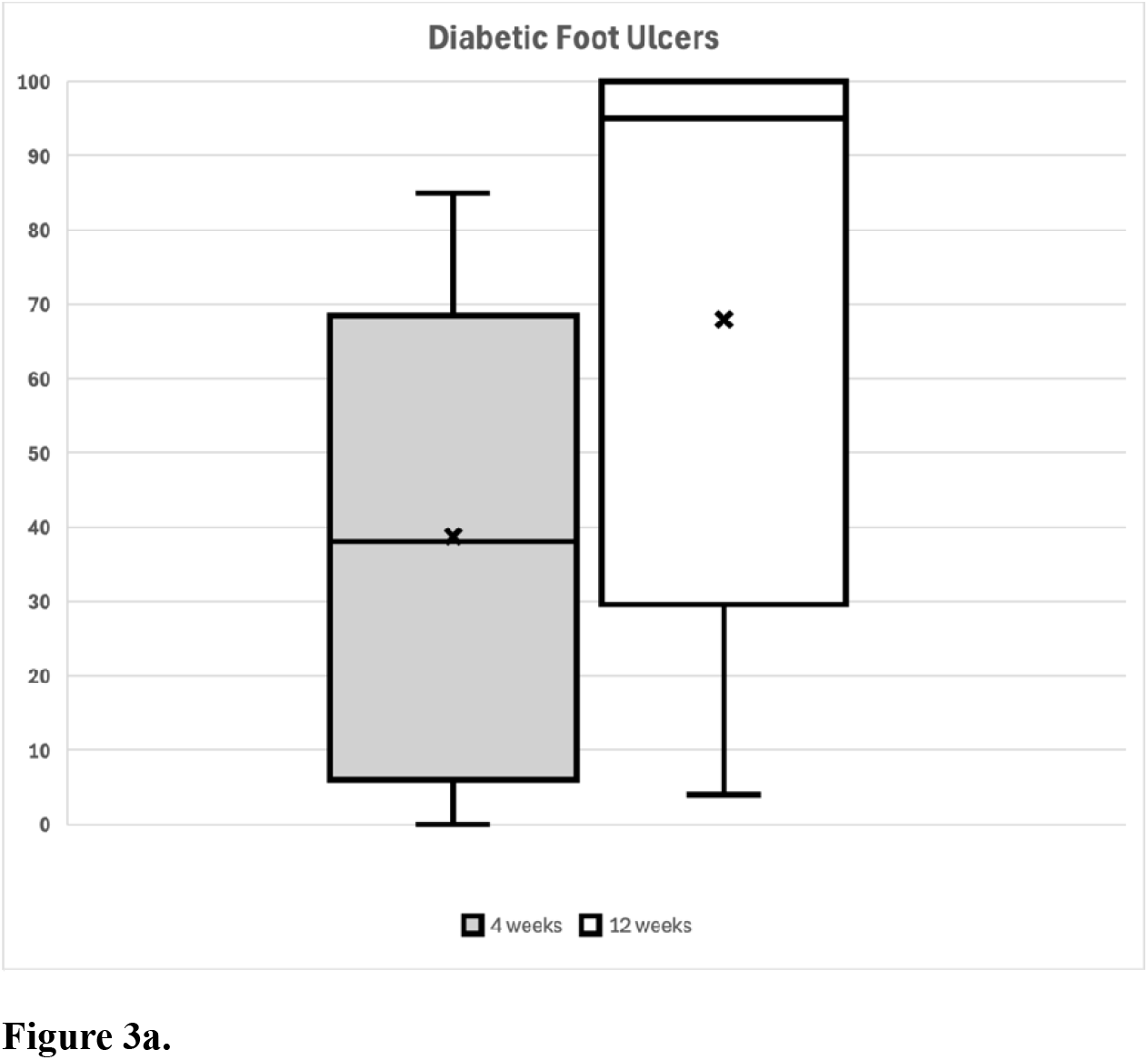
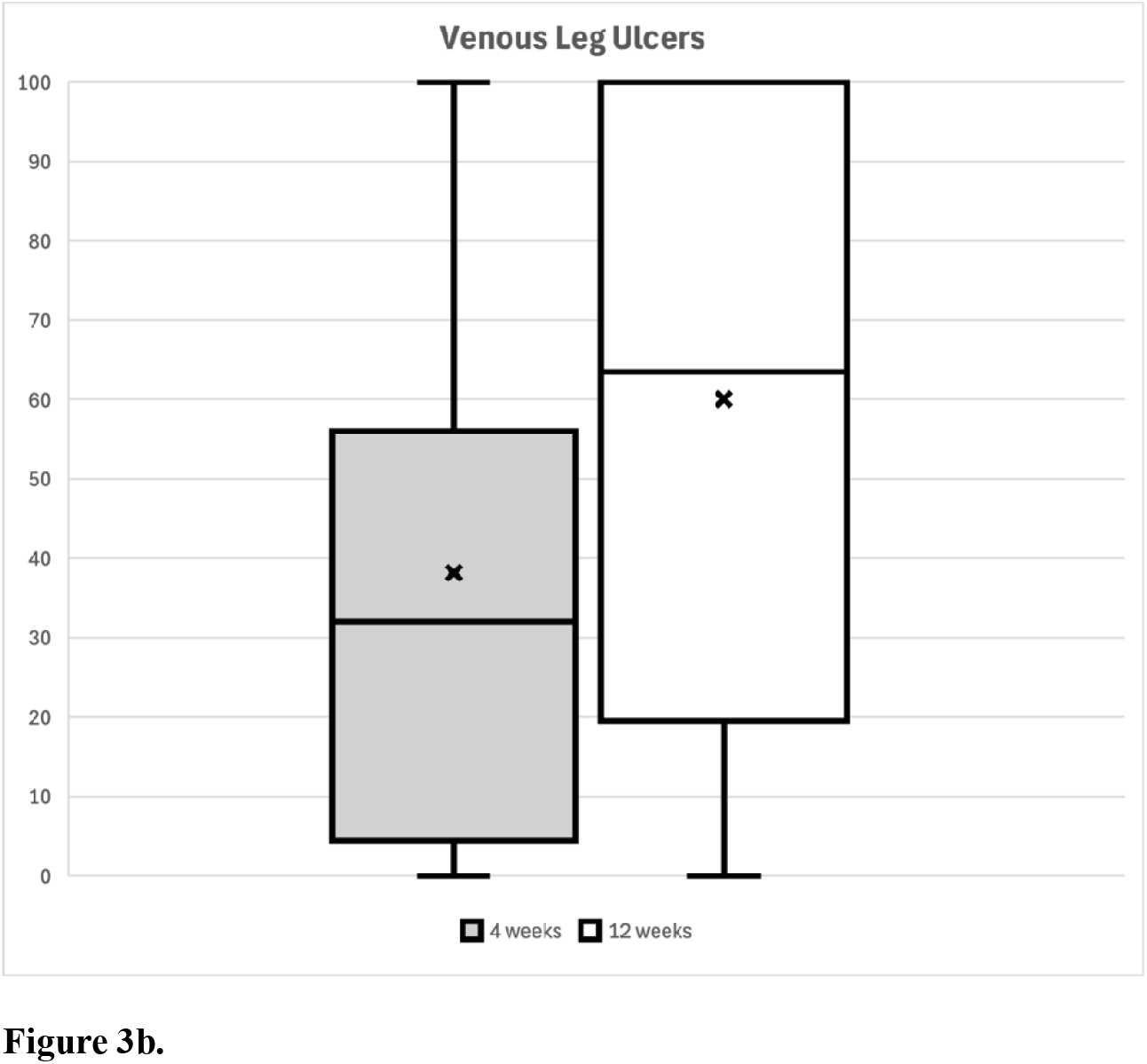
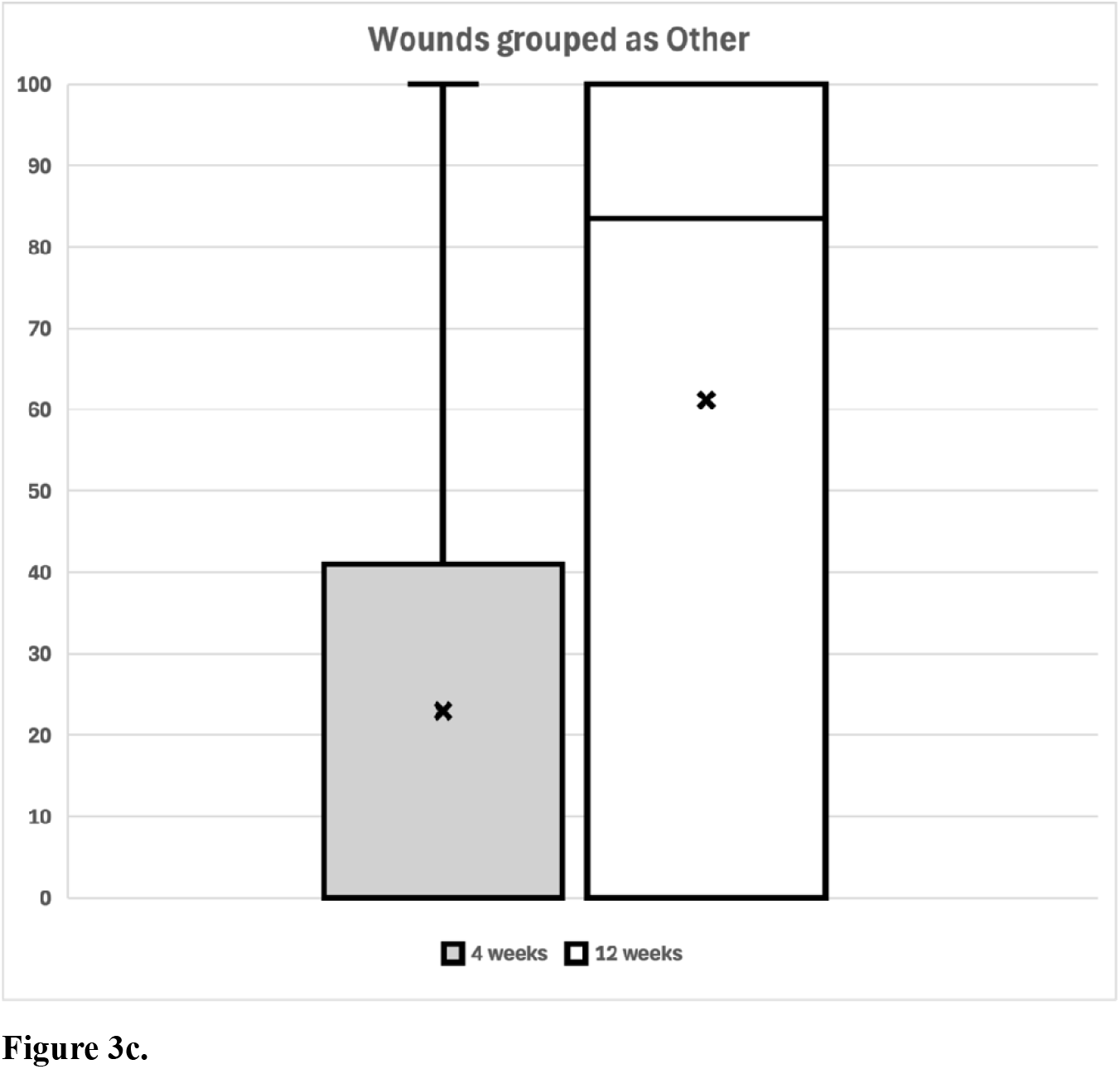
Box plots presenting the percentage area reduction grouped by wound type; diabetic foot ulcers (DFU) (3a), venous leg ulcers (VLU (3b), and wounds grouped as Other (3c), after 4 weeks of MWM treatment and at 12 weeks. X represents the mean percent area reduction for the group.

### Venous Leg Ulcers

19 patients (10 male, 9 female) with the average age of 60.9 (Median: 70) and ranging from 35-87 years were treated with MWM. The ulcer sizes ranged from 2.17 – 72.02cm^2^ with an average of 16.86cm^2^ (median: 6.08cm^2^). The age of the ulcers ranged from 13 to 32 weeks, with the average age at 19 weeks (median: 19). The 4-week PAR was 38.2% (Figure 3b). 15 of the ulcers were treated up to 12 weeks and had an average PAR of 69.4% (Figure 3b) with 8 VLUs closing completely.

### Wounds of other etiologies

16 patients (7 male, 9 female) were treated with MWM with the average age of 69.5 (Median: 70) in a range of 39-100 years. The ulcer sizes ranged from 2.12 – 42.29cm^2^ with an average of 9.55cm^2^ (median: 5.75cm^2^). The age of the ulcers ranged from 16 to 304 weeks, with the average age at 105 weeks (median: 48). The 4-week PAR was 38% (Figure 3c). 15 of the ulcers were treated up to 12 weeks and had an average PAR of 69% (Figure 3c) with 4 wounds closing completely.

### Safety Results

The 7 non-related SAEs included a death from pneumonia, myositis, hyperglycemia, and hospitalization for infection (2), fatigue, and heart failure Thirty-one AEs were not linked to the product. There were 3 AEs considered possible that included infection (2) and pain(1); 8 considered probable that included erythema (2), infection (2), exudate, irritation, yellow discharge and inflammation. The 1 AE recorded definitively linked to the investigational product was a patient that reacted with pain to the application of MWM.

### Patient Reported Outcomes

QoL, ADLs and pain assessment were collected for this trial. The data was inconclusive, and results did not consistently correlate with wound outcome or healing status.

## DISCUSSION

The healing rates presented in this study are improved over reported data. Fife, et al, reported that RCTs of uncomplicated, small ulcers of different wound types among relatively healthy subjects consistently reported mean healing rates around 40%, although these rates varied widely across trials (range: 7.7– 90.6%).^8^ Based on real-world data and RCTs, healing rates over 90% as publicly reported can be achieved only by creating extreme censoring rules, which are not likely to fall within acceptable standards of data management.^8^

In this study of non-healing refractory wounds, we enrolled patients at specialty wound clinics who had demonstrated no clinical improvement despite at least 8 weeks of standard treatment at the site and an additional 2 weeks of high-quality wound care. Subjects had failed advanced prior therapies and modalities, including gentian violet/ methylene blue foam, alginates, collagens, silver, manuka honey, cellular tissue products (CTPs), and Negative Pressure Wound Therapy (NPWT). Treatment with the MWM resulted in moving 70% of these refractory wounds towards healing.

We chose to perform a prospective real-world assessment of the MWM technology to study how this multimodal product is effective in addressing the clinical needs of high-risk patients who are managed in wound centers and other settings where wounds are treated. MWM demonstrated clinical improvement amongst subjects despite a variety of issues and comorbidities that are known to pose obstacles to healing. Evaluating a positive response marked by either progress towards or complete healing in previously unresponsive wounds among a high-risk population was deemed to have greater translational value and proof of concept of MWM’s intended technology.

This study included patients with complex medical conditions, including active smokers, poorly or uncontrolled diabetics, opioid users, Body Mass Indexes above 30kg/m^2^, and advanced age. Thirty-one of the comorbidities may have direct, adverse effects on wound healing, such as diabetes, hypertension, hyperlipidemia, coronary heart failure, and venous insufficiency. The correlation of multiple comorbidities on life expectancy has been well documented.^9^ Increased age, body mass index, opioid use and smoking status, for example, are known negative predictors of wound healing.^10,11^ Other measures such as HbA_1c_ for metabolic control, ankle brachial index for perfusion or skin temperature for inflammation are correlated with poor wound healing.^9,11-13^ However, Cho, Mattke, et al, examined the impact of comorbidities and wound characteristics as factors pertaining to healing. Their conclusion was consistent with prior research which revealed that wound level characteristics are better predictors of wound healing than patient level characteristics, such as demographic information and comorbidities.^8,11,13-15^

The current study is limited by the variability of prior wound treatment and the presence of a comparison group. However, our cohort could be compared to the prior clinical condition since the patients had wounds that were refractory to treatment from wound care specialists and had an additional 2-week of additional high-quality care. Only 12% of the enrolled cohort of patients did not progress past the screening phase because of a >30% improvement in wound size by following the study protocol. This supports the content that those remaining wounds were refractory to conventional treatment.

This study had a high number of refractory wounds which were unclassified. Of these, 8 of the 12 did not respond to treatment. Some wound etiologies may mimic others, for example, a malignancy can be misdiagnosed as a venous ulcer, therefore leading to inappropriate treatment. Not having an accurate diagnosis may negatively impact the outcome of any wound treatment, whether in a controlled study, or in an everyday clinical setting.

In this trial, regardless of wound outcome, 14 patients did not complete the treatment phase. They were withdrawn for lack of improvement or withdrew for various reasons, such as patient relocation, transportation issues, or simply no-show. Dropouts can be a problem in wound care clinical trials. A patient with more complex wounds and extensive comorbidities can find it more challenging to complete a 12-week trial with a 2-week screening period, compared to a potential subject whose health is less tenuous and wound less complex. Basic concerns such as subject reliability in presenting for weekly assessment, represent common obstacles in the recruitment of subjects who are more accurately representative of the wound population at large.

Additionally, anticipating an overall improvement in patient reported measures that would parallel wound improvement was a hopeful finding. The results correlating wound improvement as a direct impact on activities of daily living, quality of life, and pain, appeared to be inconclusive. However, in as much as these measures may be quantifiable, they are nonetheless, subjective. The presence of multiple comorbidities could also diminish the correlation that subjects’ wounds had on each of these measures, so wound improvement directly impacting these measures is potentially complicated by the effects of other existing conditions.

Despite the experience of our investigators there was wide variability in the products they each used to manage wounds of the same etiology prior to the study. Some investigators used products predominantly that contain silver, while others preferred topical antibiotics and other anti-microbials, while still others used amniotic grafts, collagens, and alginates. This example demonstrates that despite best practices, no two providers will necessarily approach the management of a wound by using the same products as their colleagues. While treatment algorithms and guidelines were previously followed by the investigators, no true standard of care was observed in terms of products applied to wounds. The wound care specialists who participated in this study chose the patients enrolled in the study because previous or current therapy was not effective. The additional 2-week run-in period prior to initiation of MWM was additional confirmation of the refractory status of the wounds studied. Once MWM was introduced through the study protocol, improvement in wounds that were previously unresponsive was evident, regardless of prior products used, wound etiology, or investigator. Besides the clinical data regarding PAR as well as weeks to closure, such improvement supports the claim that MWM demonstrated significant clinical improvement, especially in instances where prior products used had not been effective.

## Data Availability

All data produced in the present study are available upon reasonable request to the authors.

## ACKNOWLEDGEMENTS

The authors thank Vickie R. Driver, DPM, MS, FACFAS, for her guidance on and contribution to the Study Design, and Shawanda Daniel, DrPH, and Julie Primo, RN, for their contributions in support of the clinical trial.

